# Target product profiles for new diagnostics to inform strongyloidiasis control programs

**DOI:** 10.1101/2024.12.12.24318904

**Authors:** Adama Kazienga, Luc E. Coffeng, Sara Roose, Sake de Vlas, Dora Buonfrate, Salvatore Scarso, Francesca Tamarozzi, Bruno Levecke

## Abstract

**Background:** The World Health Organization calls for the development of new diagnostics to support large-scale deworming programs against strongyloidiasis. To better steer research and development (R&D) of new diagnostics, it is imperative to identify the minimal requirements that new diagnostics should meet, the so-called target product profiles (TPPs). While diagnostic TPPs exist for other major soil-transmitted helminthiases, none exist for strongyloidiasis.

**Methods:** We investigated a range of potential diagnostic TPPs using our previously developed simulation framework for the effect of imperfect diagnostics on the cost and correctness of program decisions. With this framework, we studied the minimum requirements for diagnostic performance, cost per test and sample throughput for future assays while comparing the survey costs with those of the reference Baermann method. As potential assay platforms, we considered antibody (Ab)-detecting assays, including a point-of-care lateral flow assay (LFA) and a laboratory-based Ab-ELISA. We also determined cost-efficient school-based survey designs for two currently available assays: Bordier Ab-ELISA and a prototype NIE-LFA.

**Principal findings:** Our findings highlighted that (i) specificity rather than sensitivity is a critical parameter to consider for R&D of new diagnostic methods for monitoring control programs; (ii) the requirements for diagnostic performance became less stringent with an increasing sample size and when higher risks of incorrect decision-making were accepted. When focusing on the assay formats, the LFA resulted in lower survey costs compared to the Baermann method. Ab-ELISA was cost-efficient only if the diagnostic performance was nearly perfect combined with low cost per test and high sample throughput. Of all the three assays considered here, the prototype NIE-LFA allowed for the most cost-efficient survey designs.

**Conclusion/significance:** R&D should focus on developing point-of-care assays with high specificity. The prototype NIE-LFA is a cost-efficient alternative to Baermann to support control programs for strongyloidiasis.

**Author summary:** The World Health Organization calls for the development of rapid, easy-to-use, and performant point-of-care diagnostics to follow up large-scale deworming programs against strongyloidiasis. However, there are no further recommendations regarding the required performance and cost of such new diagnostics. We performed a simulation study and a cost analysis to assess the minimum requirements in terms of diagnostic sensitivity and specificity, cost per test, and sample throughput for future assays while comparing the survey costs with those of a reference method. In addition, we determined the most cost-efficient survey designs to support control programs for strongyloidiasis applying currently available assays. Our results indicate that research & development efforts should focus on developing point-of-care assays with high specificity. Of the currently available diagnostics, a prototype of a rapid diagnostic test resulted in the lowest total survey cost, while restricting the risk of incorrect policy decisions to the minimum.

## Introduction

Strongyloidiasis is caused by the parasitic worm *Strongyloides stercoralis* and is one of the 21 neglected tropical diseases (NTDs), as classified by the World Health Organization (WHO) [1]. This parasitic roundworm is transmitted to humans through skin contact with contaminated soil, and hence belongs to the group of soil-transmitted helminths (STHs) [1–3]. As global burden, it is estimated that 600 million people worldwide are infected by *S. stercoralis*, being prevalent mainly in tropical and subtropical areas [4]. In response to this burden on public health, and after the long neglect, the WHO has now explicitly incorporated a strongyloidiasis-specific target in its 2020-2030 roadmap for STHs, namely the establishment of efficient strongyloidiasis control programs [1]. To this end, the WHO recommends preventive chemotherapy (PC) with a single oral dose of ivermectin (200 µm/kg) to entire communities in endemic areas [5].

The initiation of these large-scale deworming programs is recommended when the detected prevalence of *S. stercoralis* larvae in stool, diagnosed through Baermann method or agar plate culture [6–9], exceeds 5% (∼10% true prevalence) in a school-based survey or 10% (∼20% true prevalence) in a community-based survey. When deploying serology-based assays, the detected prevalence thresholds for initiating deworming are higher (school-based survey: 15%; community-based survey: 25%) [5].

Today, there is an unmet need for affordable and scalable diagnostics to monitor and evaluate (M&E) *S. stercoralis* infections in (suspected) endemic populations. To better steer the development of new diagnostics, it is imperative to identify the minimal requirements that such new diagnostics should meet, the so-called target product profiles (TPPs). To date, the WHO has already supported the development of diagnostic TPPs for 12 NTDs [10–19]. While a TPP exists for M&E of the major STHs control programs [13], it does not include strongyloidiasis. Here, we considered serological tests and determined the minimal requirements of diagnostic performance, sample throughput and cost per test, with the aim to support the development of a TPP for diagnostics deployed to inform strongyloidiasis control programs. In addition, we determined cost-efficient school-based survey designs for two currently available Ab-based assays: a commercially available Ab-ELISA and a prototype of a lateral flow assay (LFA).

## Methods

We assessed the minimal requirements for four attributes (sensitivity, specificity, throughput, and cost per test) for three hypothetical serological tests in three steps. In the **first step**, we applied a previously developed simulation framework [20] to determine those combinations of sensitivity and specificity that minimize the risk of incorrect program decisions in the presence of logistic constraints (the number of schools and the number of children per school that can be sampled). In the **second step**, we determined those combinations of sensitivity and specificity that also fulfilled budget constraints, where the total survey costs should not exceed that of a survey based on the Baermann method. In the **third step**, we determined the minimal requirements for sample throughput and cost per test of new diagnostics, along with their diagnostic accuracy (sensitivity and specificity), that meet both the logistic and budget constraints.

In addition, we determined the most cost-efficient survey designs when using two currently available Ab-based assays: the commercially available *Strongyloides ratti* IgG ELISA (Bordier Affinity Products, Crissier, Switzerland), and a prototype point-of-care lateral flow assay (LFA) developed by the Institute for Research in Molecular Medicine of the Universiti Sains Malaysia [6].

### Required diagnostic performance in the presence of logistic constraints

To determine those combinations of sensitivity and specificity that minimize the risk of incorrect program decisions in the presence of logistic constraints, we applied a previously developed simulation framework [20]. This framework is based on a two-stage lot-quality assurance sampling (LQAS) strategy and allows to determine the risk of incorrect program decisions when imperfect tests are deployed. The general details of this framework have been described elsewhere to inform school-based STH control programs [20]. Here, we limit ourselves to justifying the values for the eight input parameters that are required to determine the risk of incorrect decision-making (see **Table S1**). These eight input parameters can be divided into three groups. The first group represents the parameters of primary interest: test sensitivity (*se*) and specificity (*sp*). Next, we have setting-specific parameters: the program prevalence decision threshold (*T*) and the intracluster correlation coefficient (*ICC*) which represents the degree of geographical variation in prevalences. Finally, we have the framework constraints, which consist of logistic constraints and several parameters related to the risk of incorrect decisions. For the logistic constraints, we capped the maximum number of sampled schools (*n*_*schools*_) and the number of children per school (*n*_*children*_). With regard to risk of incorrect decisions, we defined the maximum allowed probability of unnecessary continuing STH control (*E*_*orertreat*_) when the true prevalence is at some value (lower limit or *LL*) under the prevalence decision threshold. Similarly, we defined the maximum allowed risk of prematurely stopping (*E*_*undertreat*_) the program when the true prevalence is at some value above the decision thresholds (upper limit or *UL*).

We let the diagnostic test sensitivity and specificity vary from 50% to 100% (with increments of 2 percentage points), resulting in 676 theoretical diagnostic tests with different diagnostic performance (26 values for sensitivity x 26 values for specificity). The program prevalence decision threshold *T* represents the true underlying prevalence that triggers distribution of ivermectin and was set at 10%, assuming that school-age children will be sampled [5]. The chosen value for the intra-cluster correlation (*ICC* = 0.0014) reflects the spatial heterogeneity in strongyloidiasis measured across 6,846 SAC from 64 schools in Ethiopia who were screened for the presence of anti-*Strongyloides* antibodies in their plasma (see **S1 Info**). We quantified the *ICC* using a generalized linear mixed model using these data. We further let the number of schools visited (*n*_*schools*_) vary from 5 to 10 with increments of 1. Assuming that at most 100 children can be sampled per school, for each combination of sensitivity and specificity we determined the minimum number of schools that would have to be sampled to achieve a sufficiently low risk of incorrect program decisions (details below). To calculate the risk of incorrect program decisions, the prevalence limits (*LL* and *UL*) surrounding *T* were arbitrarily set at *T* ± 25% × *T* (*LL* = 7.5%, *UL* = 12.5%) [20]. Then, for each combination of *se*, *sp*, and *n*_*schools*_, we determined the risk of overtreating and undertreating using Monte Carlo simulation (10,000 iterations). We kept all the combinations of sensitivity, specificity, and number of schools that satisfied the condition of ideal decision-making (*E*_*orertreatment*_ = 10% and *E*_*undertreat*_= 5%) or adequate decision-making (*E*_*orertreatment*_= 25% and *E*_*undertreat*_= 5%), as in the previous study [20].

### Required diagnostic performance in the presence of budget constraints

After having considered logistic constraints, we assessed which combinations of performance of hypothetical serological diagnostic tests (sensitivity, specificity), number of schools, and number of children of schools allow for ideal or adequate decision-making while remaining within a target budget. Here, we restricted the target budget to the total cost of a Baermann-based survey that was sufficiently powered for ideal or adequate decision-making. For each combination of diagnostic performance and sample size, we calculated the survey cost considering two alternative diagnostic assay formats for the detection of anti-*Strongyloides* antibodies (Ab): an LFA that allows for point-of-care testing of whole blood and an Ab-ELISA that can be used in a central laboratory testing approach. For the Ab-ELISA, we considered the possibility of testing either plasma or eluate from dry blood spot (DBS) samples, resulting in three implementation choices: LFA, Ab-ELISA/plasma, and Ab-ELISA/DBS. We distinguish between these three implementation choices as they have different impacts on survey costs (details below).

### Simulating the impact of diagnostic test performance and survey design

To determine the required performance of serological diagnostics, we used our simulation framework as described above, but we now allowed the number of sampled children per school to vary from 10 to 100, by increments of 2. This is because with improved diagnostic performance, generally, fewer children might be required per school. For each combination of *se*, *sp*, and *n*_*schools*_and *n*_*children*_, we performed 10,000 repeated Monte Carlo simulations and again determined the risk of overtreating and undertreating. We kept all the combinations of diagnostic performance and survey design that satisfied the conditions of ideal decision-making (*E*_*orertreatment*_ = 10% and *E*_*undertreat*_= 5%) or adequate decision-making (*E*_*orertreatment*_ = 25% and *E*_*undertreat*_ = 5%), as in the previous study [20].

For the Baerman method, we only determined which combinations of number of schools and number of children per school allowed for adequate and ideal decision-making, fixing the sensitivity and specificity at 50.0% and 98.0% as reported in literature [6]. However, we note that such high specificity can only be attained by well-trained microscopists. Since less optimal specificity requires a larger samples size (and thus more expensive surveys) [20,21], we also assessed the outcome of survey design when the specificity of the Baermann method was reduced from 98% to 93%, while keeping the sensitivity at 50%.

### Estimate the total survey costs

For each diagnostic test (Baermann and hypothetical serological tests) and survey design we calculated the total survey costs, which comprised four cost items: (i) the operational cost to inform all schools about the survey (*Cost*_*inform*_), (ii) the cost to collect and prepare all individual samples for further analysis (*Cost*_*collect*_), (iii) the cost to transport all samples to a central laboratory (*Cost*_*transport*_; Ab-ELISA only), and (iv) the cost to analyze all samples (*Cost*_*analysis*_). To estimate the total survey cost for any given survey design, we assumed that a working day consists of 8 hours. Consequently, the number of samples that can be collected and further analyzed daily is limited and varies between deployed diagnostic methods. Therefore, for each survey design and deployed diagnostic method, we determined the number of working days required to collect and further analyze all samples from the recruited children. This required number of days depends on the number of laboratory technicians in the mobile team, the number of working hours per day, the number of samples collected per hour, and the sample throughput per hour that one person can analyze (see **S2 Info)**. We considered that 4 laboratory technicians would be needed per day when deploying the Baermann method, while only 2 technicians would be needed if using the Ab-based assays (LFA, Ab-ELISA/Plasma, and Ab-ELISA/DBS). We further included the costs of various required items (see **S3 Info**) based on an itemized cost- assessment representing the real costs of a school-based survey carried out in Ecuador [6]. This itemized cost-assessment included the cost of consumables to collect one sample (*Cost*_*collect*_), the cost to prepare one sample (*Cost*_*prep*_), the cost to test one sample (*Cost*_*test*_), the daily salary for every technician and nurse in the mobile team (*Cost*_*tech*_), the daily salary for local helper (*Cost*_*helper*_, only required when deploying the Baermann method) and the daily cost of car rental, petrol, and driver wages (*Costdrirers*). Note that we did not consider costs for establishing and maintaining laboratory infrastructures. We further assumed that all work takes place on regular working days and that the team does not take any breaks during processing. Further details on how we estimated the total survey costs can be found in **S2 Info.**

### Required diagnostic performance to not exceed the total cost of Baermann-based survey

After we determined the total survey cost for each combination of diagnostic performance and survey design, we set a benchmark for the total survey cost for new hypothetical serological tests, using Baermann as the reference. For this, we considered those combinations of *n*_*schools*_and *n*_*children*_that allowed a Baermann-based survey to result in adequate and ideal decision-making for the lowest total survey cost. Given this benchmark cost, we identified for each implementation scenario (LFA, Ab-ELISA/plasma, and Ab-ELISA/DBS) those combinations of diagnostic sensitivity and specificity that were logistically feasible and resulted in a total survey cost that did not exceed the benchmark cost.

### Minimal requirements for sample throughput and cost per test

We previously showed that higher diagnostic performance allows for a reduction of sample size for the same level of program decision-making (adequate or ideal), thus allowing for less stringent requirements in terms of sample throughput and cost per test [20]. We therefore explored the minimal requirements for sample throughput and cost per test such that the total survey cost does not exceed that of a survey based on the Baermann method (benchmark cost). To this end, we repeated the total survey cost analysis for the three implementation scenarios while varying the sample throughput (5, 10, 20, and 40 samples per hour) and cost per test (0.20 EUR to 15 EUR, with 0.20 EUR increment). Finally, we determined those combinations of diagnostic performance, sample throughput and cost per test that met both the logistic and budget constraints.

### Most cost-efficient survey design for existing serological assays

We determined the most cost-efficient survey designs for two existing serological assays, including a commercially available Ab-ELISA and a prototype LFA. For the Ab-ELISA, we based our assumptions about sensitivity (79%) and specificity (93%) on the commercially available *Strongyloides ratti* IgG ELISA (Bordier Ab-ELISA, Bordier Affinity Products, Crissier, Switzerland), based on crude antigens of the parasite *Strongyloides ratti*. For the prototype LFA, we considered a cassette-format immunochromatographic rapid diagnostic test developed by the Institute for Research in Molecular Medicine of the Universiti Sains Malaysia. This LFA is based on the recombinant antigen NIE (NIE-LFA) [22]. To determine the most cost-efficient survey design for these assays, we calculated the total survey costs for each combination of survey design (*n*_*schools*_ x *n*_*children*_) as described in **S2 Info**, while fixing the sensitivity and specificity at 79.0% and 93.0%, respectively [6]. These tests are assumed to only differ in terms of cost to test one sample, cost to prepare sample, cost to test one sample and the number of samples that can be analyzed by one person in one hour (see **S2 Info**). The most cost-efficient survey design is the one (*n*_*school*_x *n*_*children*_) that minimized the total survey cost while attaining an adequate and an ideal program decision-making.

## Results

### Required diagnostic performance in the presence of logistic constraints

Given the presence of logistic constraints, we investigated the combinations of sensitivity and specificity that minimize the risk of incorrect programs decisions. **Fig 1** represents the required diagnostic performance (minimum sensitivity and specificity) for adequate (**Fig 1A**) and ideal (**Fig 1B**) decision-making when sampling 100 children per school (the assumed maximum logistically feasible number) across five to 10 schools. Three important points can be noted in this figure. First, requirements are more stringent for specificity than for sensitivity. For example, specificity cannot be below 72% for adequate decision-making (**Fig 1A**) and 84% for ideal decision-making (**Fig 1B**), whereas sensitivity could be as low as 50% in both cases. Second, the requirements for sensitivity and specificity are inversely correlated. For example, when sampling seven schools with a theoretical test with a specificity of 98%, the sensitivity should be at least 50% to allow adequate decision-making, while when deploying a theoretical test with a specificity of 84%, the sensitivity should be at least 92%. Third, the requirements for diagnostic performance become less stringent as a function of an increasing number of schools (thus more children are screened) and increasing risk of incorrect decision-making. When focusing on adequate program decision-making and fixing the specificity at 96% (**Fig 1A**), the required sensitivity was 58% when 7 schools were sampled, while it was 50% when 10 schools were included. When now focusing on ideal program decision-making (**Fig 1B**) while fixing the specificity at 96% and the number of schools at 7 (**Fig 1B**), the required sensitivity was 78% (instead of 58% for adequate decision making)

**Fig 1.**
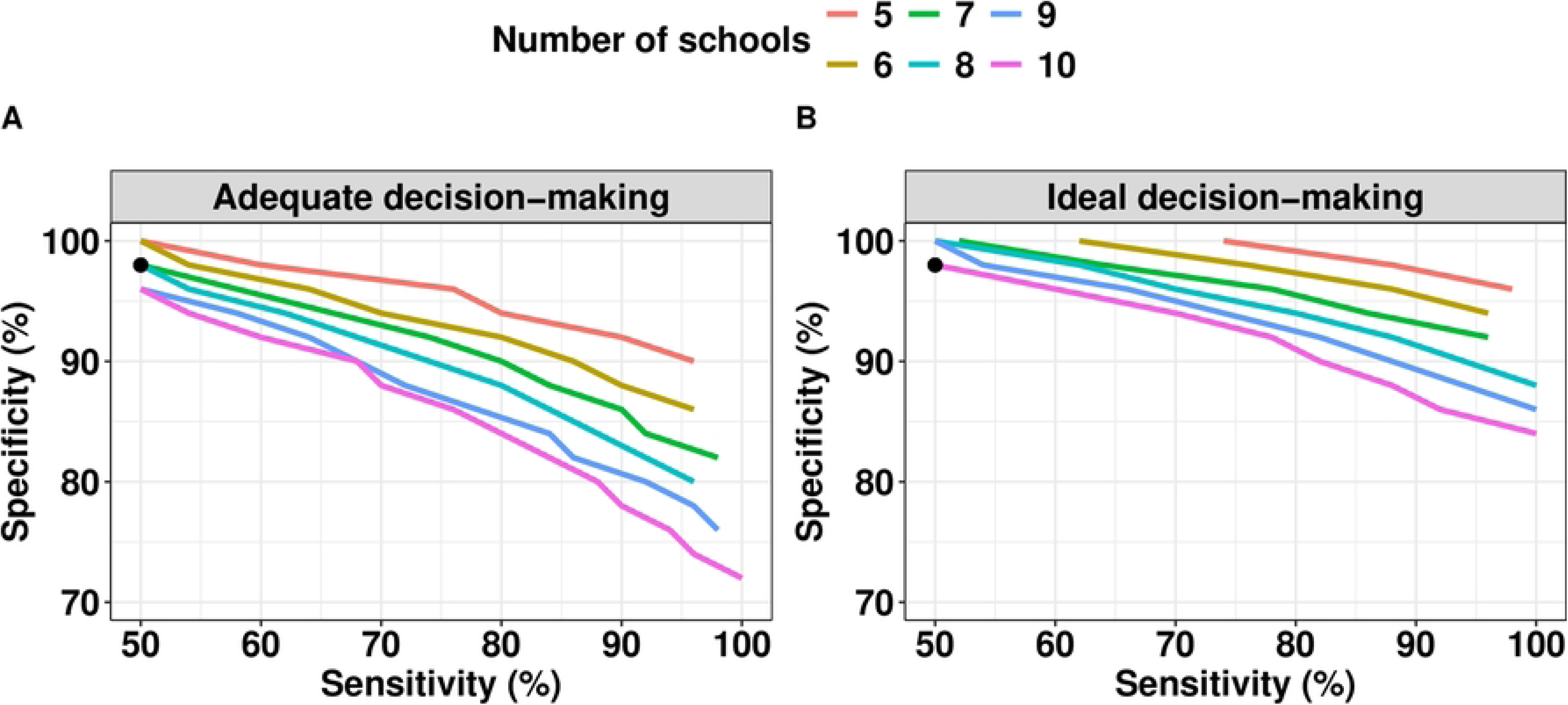
Required sensitivity and specificity when making adequate and ideal program decisions in the presence of logistic constraints. The figure indicates the required sensitivity and specificity for adequate (*E*_*undertreatment*_ = 5% and *E*_*orertreatment*_ = 25%; **Panel A**) and ideal decision-making (*E*_*undertreatment*_ = 5% and *E*_*orertreatment*_ = 10%; **Panel B**) when sampling 100 children per school for different number of schools (colored lines). The black dot represents the diagnostic performance of the Baermann method (sensitivity = 50.0% and specificity = 98.0%).

### Required diagnostic performance in the presence of budget constraints

While we have considered logistic constraints so far, we further explored those combinations of sensitivity and specificity that allowed for adequate and ideal decision-making in the presence of budget constraints. For this, we explored three implementation choices (LFA, Ab-ELISA/plasma and Ab-ELISA/DBS) while not allowing the total survey cost to exceed the cost of a Baerman-based survey. For adequate decision-making, the most cost-efficient survey design for Baerman-based survey involved a total of 644 children (= *n*_*school*_ × *n*_*child*_ = 7 × 92), resulting in a total survey cost of 9,410 EUR. As soon as 41 children test positive (= decision cutoff *c*), one could already stop screening and initiate a strongyloidiasis control program. For ideal decision-making, the most cost-efficient design involved a total of 980 children (= *n*_*schools*_ × *n*_*children*_ = 10 × 98), resulting in an estimated total survey cost of 13,968 EUR. For this level of decision-making, 65 positive test results would trigger large-scale deworming. If the assumed specificity of Baermann was 93% instead of 98% because of insufficiently trained laboratory technicians, the required sample size was beyond what was considered logistically feasible (adequate: 13 schools x 96 children per school; ideal: 20 schools x 96 children), resulting in a total cost of 17,625 EUR (adequate) and 27,103 EUR (ideal). We will therefore base the benchmark for total survey costs on the assumption that the Baermann method is performed by well-trained technicians (i.e., specificity of 98%).

After defining the cost benchmark, we continued identifying those combinations of sensitivity and specificity of hypothetical new serological tests that allowed for a survey that was not more expensive than the most cost-efficient Baermann-based survey design. **Table 1** presents the required diagnostic performance (sensitivity and specificity) for adequate and ideal decision-making when deploying an LFA and Ab-ELISA/Plasma in the presence of both logistic and budget constraints. When focusing on LFA, two crucial points can be made. First, this table indicates that when an LFA format is applied, the test performance can take a wide range of values for sensitivity and specificity, the two parameters again being inversely correlated. Second, when looking more closely into the absolute values, they represent all possible combinations of sensitivity and specificity illustrated in **Fig 1** (all combinations above the purple line). In other words, even when the sampling effort is maximized (10 schools and 100 children per school), the total survey costs never exceeded that of a Baermann-based survey, highlighting that the budget constraints had no impact on the requirements for the diagnostic performance of LFA.

**Table 1.** Required specificity and sensitivity for adequate and ideal decision-making when deploying LFA and Ab-ELISA/Plasma in the presence of both logistic and budget constraints. The table summarizes the sensitivity and specificity for the LFA and Ab-ELISA/Plasma to achieve adequate (*E*_*undertreatment*_ = 5% and *E*_*orertreatment*_ = 25%) and ideal program decision-making (*E*_*undertreatment*_ = 5% and *E*_*orertreatment*_ = 10%). Each of these combinations result in a total survey cost that does not exceed that of a Baermann-based survey (assuming a specificity of 98% and sensitivity of 50% for the Baermann method). Ab-ELISA/Plasma: Ab-ELISA assays that are based on plasma. Note that the blank cells indicate that there were no combinations of sensitivity and specificity that resulted in sufficiently low risk of incorrect decision-making.

| Specificity (%) | Minimum sensitivity LFA (%) |  | Minimum sensitivity Ab-ELISA/Plasma (%) |  |
| --- | --- | --- | --- | --- |
|  | Adequate decision-making | Ideal decision-making | Adequate decision-making | Ideal decision-making |
| 100 | 50 | 50 | 80 | 82 |
| 98 | 50 | 50 | 94 | 96 |
| 96 | 50 | 60 |  |  |
| 94 | 54 | 70 |  |  |
| 92 | 60 | 78 |  |  |
| 90 | 68 | 82 |  |  |
| 88 | 70 | 88 |  |  |
| 86 | 76 | 92 |  |  |
| 84 | 80 | 100 |  |  |
| 82 | 84 |  |  |  |
| 80 | 88 |  |  |  |
| 78 | 90 |  |  |  |
| 76 | 94 |  |  |  |
| 74 | 96 |  |  |  |
| 72 | 100 |  |  |  |

For Ab-ELISA-based surveys, which are more expensive ( see **S2 Info**), the impact of budget constraints on the requirements for diagnostic performance was more pronounced, resulting in fewer and more stringent diagnostic performance options. For an Ab-ELISA-based survey to be as cost-efficient as a Baermann-based survey for adequate decision-making, its required sensitivity should be at least 80%, while its specificity cannot be below 98% (**Table 1**). Additionally, the required number of schools and children that need to be sampled was lower for an Ab-ELISA-based surveys compared to LFA (5 schools *vs.* 10 schools and 100 children per school).

### Minimal requirements for sample throughput and cost per test

We then looked at how better diagnostic performance allows for less stringent sample throughput and cost per test for all logistically feasible surveys. When focusing on LFA (**Fig 2A** and **2B)**, we note two important aspects. First, results confirm that improving the sensitivity and specificity of a test allowed for higher costs per test (shift from dark green to red colored tiles), with the highest possible cost per test for a LFA with 100% sensitivity and specificity. As illustrated in **Fig S1**, improving the diagnostic performance required smaller sample sizes for the same level of program decision-making, thus allowing to have higher cost per test for the same total survey cost. Second, by comparing **Figs 2A** and **2B**, we can see that the maximum cost per test was very similar for tests with lower (5 samples per hour) and higher throughput (40 samples per hour). For example, when fixing diagnostic performance at 92% specificity and 74% sensitivity, the maximum cost per test equaled 6.4 EUR when five samples per hour were processed, and 7.6 EUR when 40 samples per hour can be processed. As such, improving the diagnostic performance allows for a relatively higher maximum cost per test than improving the sample throughput.

**Fig 2.**
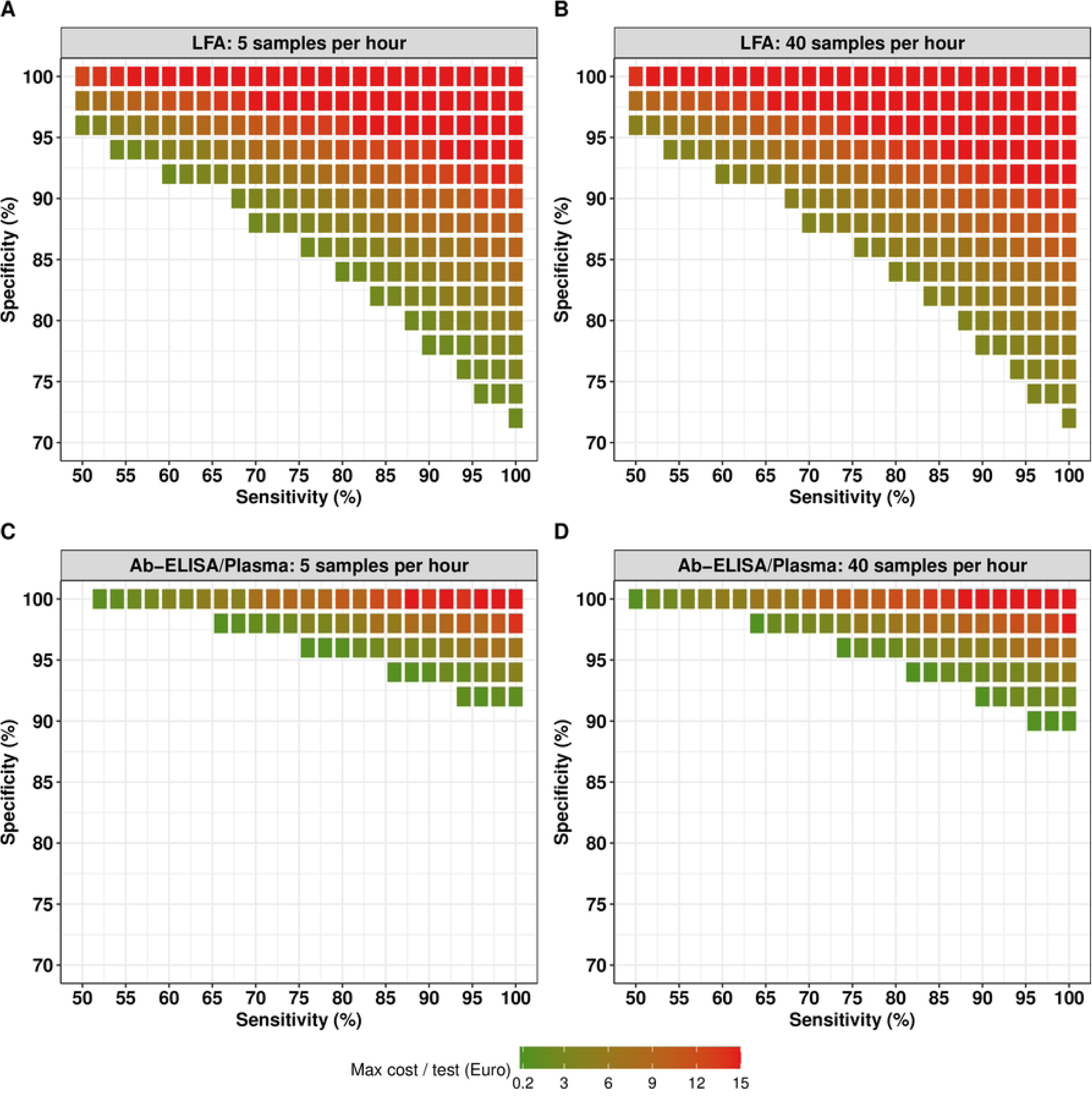
The maximum cost per test for an LFA and Ab-ELISA/Plasma for adequate decision making. This figure presents the maximum cost per test for all possible combinations of sensitivity and specificity, sample throughput (number of samples that can be analyzed by one technician in one hour) that allowed for adequate (*E*_*undertreatment*_ = 5% and *E*_*orertreatment*_= 25%) decision-making for an LFA (**Panel A:** sample throughput of five samples per hour; **Panel B:** sample throughput of 40 samples per hour) and Ab-ELISA format (**Panel C:** sample throughput of five samples per hour; **Panel D:** sample throughput of 40 samples per hour). Note that the maximum cost per test is the true maximum cost per test that could work in at least one logistically feasible survey design that is not more expensive than the benchmark cost based on the Baermann method. Absence of tiles indicates that the combination of sensitivity and specificity was inadequate for decision-making, given the logistic and budget constraints.

When comparing these findings for LFA (**Figs 2A** and **2B**) with those for Ab- ELISA/Plasma (**Figs 2C** and **2D**), two important differences can be noticed. First, as expected from the performance options in presence of budget constraints (**Table 1**; LFA: **Fig 2A** *vs.* Ab-ELSA: **Fig 2C**), there are fewer and more stringent diagnostic performance options for Ab-ELISA (fewer colored tiles in the figure). Second, the requirements for the cost per test are stricter for Ab-ELISA (fewer red tiles), indicating that the cost per test should be relatively cheap. For example, if we focus on a throughput of five samples per hour, the maximum cost for test with a specificity of 100% and sensitivity of 52% was 13.6 EUR when deployed as LFA format, while this was 1.0 EUR when deployed as an Ab-ELSA format.

We next focus on the maximum cost per test for an LFA and Ab-ELISA/Plasma across different combinations of diagnostic performance and sample throughput for ideal (instead of adequate) decision-making (**Fig 3**). Overall, we noted two aspects. First, for ideal decision-making (**Fig 3**), patterns in maximum cost per test (as function of sensitivity, specificity and throughput) are essentially the same as for adequate decision-making (**Fig 2**). However, diagnostic performance requirements are more stringent, meaning there are fewer options. Next, given that the benchmark cost for ideal decision-making was set to be higher than for adequate decision-making, the maximum cost per test for a given diagnostic performance was actually very similar between the two levels of certainty in decision-making.

**Fig 3.**
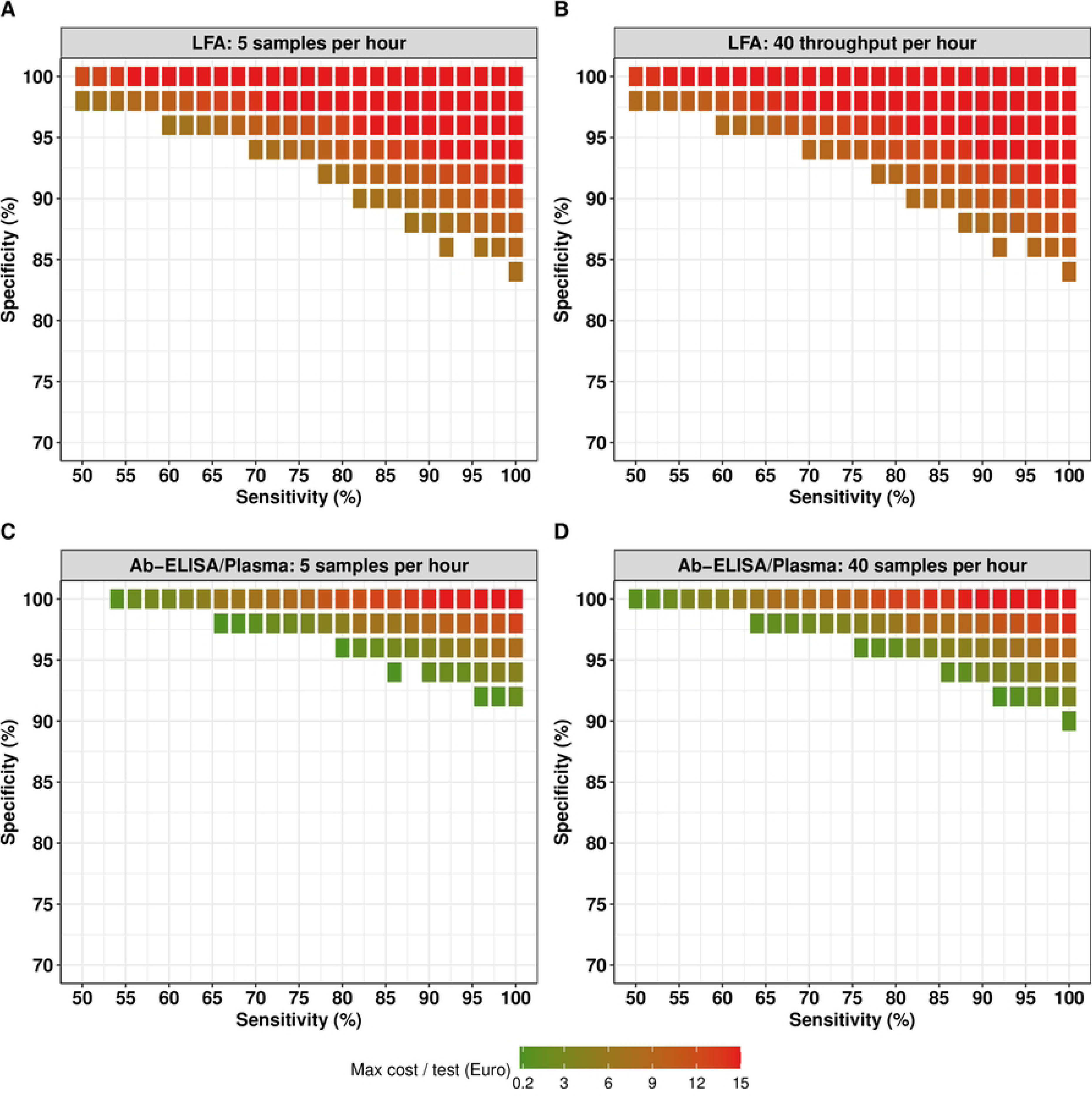
The maximum cost per test for an LFA and Ab-ELISA/Plasma for ideal decision making. This figure represents all possible combinations of sensitivity and specificity, sample throughput (number of samples that can be analyzed by one technician in one hour), and the maximum cost per test (in EUR) that allowed for adequate (*E*_*undertreatment*_ = 5% and *E*_*orertreatment*_ = 10%) decision-making for an LFA (**Panel A:** sample throughput of five samples per hour; **Panel B:** sample throughput of 40 samples per hour) and Ab-ELISA format (**Panel C:** sample throughput of five samples per hour; **Panel D:** sample throughput of 40 samples per hour). Note that the maximum cost per test is the true maximum cost per test that could work in at least one logistically feasible survey design that is not more expensive than the benchmark cost based on the Baermann method. Absence of tiles indicates that the combination of sensitivity and specificity was inadequate for decision-making, given the logistic and budget constraints.

### Most cost-efficient survey design for existing Ab-based assays

We finally determined the most cost-efficient survey designs when deploying two currently available Ab-based assays. **Table 2** summarizes the most cost-efficient survey designs that allowed for adequate and ideal decision-making for these two tests when assuming the diagnostic performance reported by Tamarozzi et al. [6] (sensitivity = 79% and specificity = 93%). Compared to the Baermann method, two important differences can be noted for these Ab-based assays. First, the required sample size for the survey is slightly smaller when deploying Ab-based assays. For instance, when considering adequate decision-making, a total of 644 children (= *n*_*school*_ × *n*_*child*_ = 7 × 92) need to be screened when using the Baermann method. This required sample size decreased to 564 children (= *n*_*school*_ × *n*_*child*_ = 6 × 94) when deploying Ab-based assays. Second, despite this smaller sample size, the surveys were more expensive when deploying Ab-based assays compared to the cost of a cost-efficient Baermann-based survey. For instance, when deploying the Bordier Ab-ELISA, the total survey costs were 81% times higher when applied on plasma (Bordier Ab- ELISA/Plasma) and 82% times higher when applied on DBS (Bordier Ab-ELISA/DBS). In contrast, deploying LFA resulted in cheaper surveys compared to the cost of a cost- efficient Baermann-based survey. For example, for adequate decision-making, the total survey costs for NIE-LFA were 52% times lower than that of surveys based on the Baermann method. If these Ab-based tests are implemented (6 schools and 94 children per school), 76 positive test results would trigger large-scale deworming programs for adequate decision making and 123 for ideal decision-making (9 schools and 98 children per school).

**Table 2.** The most cost-efficient survey design to initiate large-scale deworming against strongyloidiasis when deploying Baermann method and existing Ab-based assays. . This table represents the required sample size (*n*_*scchool*_ × *n*_*children*_), the decision cutoff *c*, and the total survey cost to assess whether the true prevalence of strongyloidiasis is under or above 10% for adequate and ideal decision-making. Bordier Ab-ELISA represents a commercially available *Strongyloides ratti* IgG ELISA (Bordier Affinity Products, Crissier, Switzerland) applied on either plasma (Bordier Ab-ELISA/plasma) or dried blood samples (Bordier Ab-ELISA/DBS). NIE-LFA is a prototype (research use only) lateral flow assay (LFA) developed by the Institute for Research in Molecular Medicine of the Universiti Sains Malaysia. This LFA is based on the recombinant antigen NIE.

| Method | Sensitivity | Specificity | $n_{schools}$ | $n_{children}$ | Decision cutoff ($c$) | Total survey cost (EUR) |
| --- | --- | --- | --- | --- | --- | --- |
| **Adequate decision making** |  |  |  |  |  |  |
| Baermann method | 0.50 | 0.98 | 7 | 92 | 41 | 9,410 |
| Bordier Ab-ELISA/Plasma | 0.79 | 0.93 | 6 | 94 | 76 | 17,067 |
| Bordier Ab-ELISA/DBS | 0.79 | 0.93 | 6 | 94 | 76 | 17,197 |
| NIE-LFA | 0.79 | 0.93 | 6 | 94 | 76 | 4,949 |
| **Ideal decision making** |  |  |  |  |  |  |
| Baermann method | 0.50 | 0.98 | 10 | 98 | 65 | 13,968 |
| Bordier Ab-ELISA/Plasma | 0.79 | 0.93 | 9 | 98 | 123 | 26,391 |
| Bordier Ab-ELISA/DBS | 0.79 | 0.93 | 9 | 98 | 123 | 26,448 |
| NIE-LFA | 0.79 | 0.93 | 9 | 98 | 123 | 7,424 |

## Discussion

The WHO calls for the development of rapid, easy-to-use, and performant point-of-care diagnostics for M&E of large-scale deworming programs [1]. While a TPP exists for M&E of STH control programs, it does not include strongyloidiasis. Therefore, we determined the minimal requirements of diagnostics for strongyloidiasis in terms of diagnostic performance (sensitivity and specificity), sample throughput and cost per test for diagnosing strongyloidiasis, with the aim to further support the development of a diagnostic TPPs for this NTD.

In the presence of logistic constraints (the number of schools that can be visited per day and the number of children per school are limited), we found that sensitivity and specificity are inversely correlated, and that specificity is critical to consider when developing a diagnostic assay to be deployed in population-based surveys. Indeed, depending on the acceptable risk of making a wrong program decision, specificity should be at least 72% for adequate and 84% for ideal or even higher for both (e.g. sensitivity is less than 100%). For the sensitivity, it should be at 50% for both program decision-making (adequate and ideal) or even higher in case the specificity is less than 100% (**Fig 1**). This need for highly specific diagnostics aligns with the results of previous studies [20,21] and the recently published WHO TPPs for diagnostics to be deployed in M&E of large-scale deworming programs for NTDs such as lymphatic filariasis, onchocerciasis, and schistosomiasis [11–14]. In each of these TPPs, the minimum required specificity is 94%, while the minimum required sensitivity was set to 60%, which is roughly comparable to our recommendations for strongyloidiasis diagnostics.

When also considering budget constraints and exploring different implementation choices (LFA, Ab-ELISA/plasma, and Ab-ELISA/DBS) and scenarios of sample throughput and cost per test, our results indicated that a LFA holds the most promise as a diagnostic format to take cost-efficient program decisions. Implementing an LFA allowed for a lower total survey costs compared to a Baermann-based survey, even when its diagnostic performance is less optimal (lower specificity compared to Baermann) and the cost per test is higher. In contrast, surveys based on Ab-ELISA turned out to be more expensive compared to Baermann method, unless a nearly perfect diagnostic performance, low cost per test, and high sample throughput was reached. This can be explained by the additional cost of needing to prepare the samples for testing (Ab-ELISA/plasma: 9.27 EUR per sample and Ab-ELISA/DBS: 11.49 EUR per sample) and the cost per test (10.79 EUR). A potential cost-saving strategy would be using the same samples to test for multiple NTDs (or other diseases) at the same time, and thus spreading the costs of the sample preparation step over multiple disease control programs. This would also align with the WHO vision for integrated disease surveillance and control, offering a streamlined approach to tackling NTDs alongside other infectious diseases [1,13].

These results are particularly welcomed since, recently, a number of immunochromatographic rapid tests for the diagnosis of strongyloidiasis have been developed, most of which based on the recombinant antigens NIE and SsIR, [23] Currently, none of these tests are commercially available, and they have been evaluated mostly in laboratory-based studies [24–28]. However, their performance seems promising in a number of studies (complying with the requirements defined here, **Table 1**), importantly including two field-based projects [6,24,29]. These findings, although requiring confirmation and validation in larger cohorts and different settings, support further R&D of this type of assays for strongyloidiasis control programs. R&D should carefully consider the physical characteristics of the LFA assay, such as cassette *vs.* dipstick format, as these could impact the test’s method protocol, diagnostic performance, cost per test, and material disposal requirements.

Finally, we defined cost-efficient survey designs for the control of strongyloidiasis when deploying Baermann and two existing Ab-based tests: the Bordier Ab-ELISA and NIE- LFA. When deploying the reference Baermann method in a school-based survey, we estimated the need to sample seven schools (92 children per schools) and 10 schools (98 children per school) for adequate and ideal decision-making, respectively. These survey designs minimize the total survey cost, while ensuring reliable decision making (adequate: 9,410 EUR; ideal: 13,968 EUR). When deploying the NIE-LFA, the total survey costs could be further reduced (adequate decision making 4,949 EUR and ideal decision making 7,424 EUR), making it a cost-efficient alternative to Baermann. On the contrary, the Bordier Ab-ELISA turned out to be more resource demanding than LFA, due to the high costs per test (Bordier Ab-ELISA: 10.79 EUR *vs.* NIE-LFA: 2.09 EUR) and the costs to prepare the samples for laboratory testing (Ab-ELISA/plasma: 9.27 EUR and Ab-ELISA/DBS: 11.49 EUR). Another reason to recommend an (NIE-)LFA over the Baermann method, is the feasibility of implementing this kind of diagnostic test. Indeed, we noted that the Baermann method needs adequate training to ensure the high specificity (98%) reported in the ESTRELLA study [6], while the training requirement to ensure accurate test performance is minimal for the (NIE-)LFA. Using a less experienced team when deploying Baermann may result in a reduced specificity, which in turn will increase the sample size (and total survey cost) required to make adequate or ideal decision making. Note that all our recommended survey designs aim to be no more costly than a well-trained team of experienced technicians.

Therefore, in case we accept that the benchmark cost does not necessarily represent a team of experienced technicians, then such a cost benchmark would allow the requirements for new diagnostics to be less stringent (lower performance, lower sample throughput, and higher cost per test).

As a limitation of our study, it is important to note that the estimated costs were based on data obtained from a school-based survey carried out in Ecuador, and hence the reported values should not be interpreted as absolute. Despite of this, we are confident that our general conclusions are robust to context-dependent changes in cost components. A major strength of our simulation framework is therefore that assumptions about these itemized costs can be easily adapted to represent any setting.

## Conclusions

We provided insights into the required diagnostic performance, cost per test and sample throughput that may steer R&D choices for new diagnostics to inform strongyloidiasis control programs. When focusing on Ab-based assays, our results indicate that R&D should focus on the development of point-of-care diagnostics or laboratory-based formats that allow for testing multiple NTDs on the same sample. Of all current diagnostic tests, the NIE-LFA was a cost-efficient alternative to Baermann to make program decisions, encouraging its further development for commercialization.

## Data Availability

All relevant data are within the manuscript and its supporting information files

## Acknowledgments

We are grateful to our partner institutions in Ecuador: Mariella Anselmi, Cintia Caicedo and Rosanna Prandi from the Centre for Community Epidemiology and Tropical Medicine (CECOMET) and Yosselin Vicuña from the Universidad Central del Ecuador, Quito, Ecuador, for providing support for completing the itemized costs of materials used in the current study.

## Supporting information

**S1 Table. Values of the required parameters to assess the risk of incorrect decision-making when currently available test platforms are deployed.**

**S1 Info. Quantification of spatial heterogeneity in strongyloidiasis**

**S2 Info. Estimation of the total survey cost**

**S3 Info. Itemized cost-assessment**

**Fig S1. The impact of diagnostic performance on maximum cost per test and required sample size.** This figure represents all possible combinations of sensitivity and specificity and the maximum cost per test (in EUR) (**Panel A:** LFA and **Panel C:** Ab-ELISA/Plasma) and the required sample size (**Panel B:** LFA and **Panel D:** Ab-ELISA/Plasma) that allowed for adequate (*E*_*undertreatment*_ = 5% and *E*_*orertreatment*_ = 25%) decision-making. Note that the maximum cost per test is the true maximum cost per test that could work in at least one logistically feasible survey design that is not more expensive than the benchmark cost based on the Baermann method. Also, the required sample size corresponds to the minimum required number of schools and children per school that need to be sampled for each combination of sensitivity and specificity. Absence of tiles indicates that the combination of sensitivity and specificity was inadequate for decision-making, given the logistic and budget constraints.

